# Demographic trends and forecasts of alcohol-associated liver disease in the United States, 2008-2030

**DOI:** 10.64898/2026.05.09.26352799

**Authors:** Alex Viguerie, Elisa Iacomini, Maria R. D’Orsogna

## Abstract

Alcohol-associated liver disease (ALD) has been steadily increasing in the United States for many years, as attested by increases in ALD deaths and liver transplant demand. Direct measurement of ALD incidence is challenging as diagnosis often occurs late (or not at all). This study employs a demographically-aware backcalculation method, based on mortality data, to reconstruct latent, age-structured ALD risk and incidence trends in the US population from 2008 to 2022 and uses this information to forecast future ALD trends through 2030. We find that ALD incidence has risen steadily since 2008, with a sharp increase during the 2020 COVID-19 pandemic, and that the average age at onset has also increased over time, with demographic factors playing a substantial role. While our forecasts suggest a continuation of the pre-2020 growth in ALD incidence for most age and sex groups, we also predict marked increases among younger men, a generational shift toward older age cohorts, and substantial rises among older females. Most concerning, between 2022 and 2030, incidence is expected to double among younger men and older females and by 2030 the number of new male ALD cases is projected to be more than twice that of females for all age groups. Our results provide a clearer understanding of evolving ALD trends, highlighting the role of demographic and birth cohort effects. We underscore the urgent need for targeted interventions, particularly among younger men, to reduce ALD-related behaviors and future burden.

## 1 Introduction

Alcohol-related diseases constitute a major public health burden in the United States. Sustained alcohol use can lead to liver disease, cardiovascular dysfunction, neurological decline, and increased cancer risks [1, 2]. Depending on intensity and duration, alcohol consumption may reduce life expectancy by several decades [3]. Recent CDC estimates indicate an average loss of 29 years per alcohol-attributable death [4]; one in ten deaths among working-age adults is due to excessive drinking [5]. Alcohol-induced deaths have increased over time across all demographic groups, rising sharply during the COVID-19 pandemic and remaining elevated despite modest post-pandemic declines [6]. In 2024, alcohol was the primary cause of nearly 47,000 deaths [7]; when including chronic diseases, injuries, and conditions partly related to alcohol, the total is about 178,000 annual deaths, based on 2021 data [3].

Many fatal alcohol-related diseases develop gradually and reflect the cumulative effects of prolonged or heavy alcohol use; onset and progression are influenced by genetics, comorbidities, drinking patterns, and age. The latter plays a particularly important role [8, 9]. For example, liver injury due to heavy drinking typically begins with steatosis (fatty liver) which can occur at any age, but while younger livers are more likely to recover, age-related decline limits regeneration in older adults [10, 11, 12]. Age is also a major determinant in the progression of liver disease, from steatosis to fibrosis, cirrhosis, and liver failure [13, 14]. Similarly, alcohol use can damage heart tissue, with age-related declines in cardiac function and repair accelerating the onset and severity of cardiomyopathy [15].

Given age’s central role in alcohol-related disease, age-stratified analyses can help identify high-risk cohorts and guide targeted intervention. Quantifying age-specific risk and disease onset however is challenging since behavioral patterns vary across individuals, reliable biomarkers are lacking, and comorbidities affect disease progression [16]. The relationship between age and disease onset is further confounded by birth cohort effects that are independent of age, possibly reflecting alcohol-related behavioral shifts [17, 18, 19, 20]. Incomplete tracking and under-reporting further complicate accurate estimations.

In this study, we aim to determine the age-specific risk and incidence of alcohol-associated liver disease (ALD) in the United States and to forecast future trends through 2030, using data from 2008 to 2024. Incidence refers to the number of new ALD cases in a given year, whereas risk is the probability that an individual will develop ALD within a given year. We focus on ALD because it is a major contributor to alcohol-related morbidity and has well-defined clinical diagnostic and reporting criteria [21, 22, 23]. Furthermore, since ALD develops over many years, even decades, its incidence may reflect complex interactions between time, age, and birth cohort which may not be necessarily apparent from mortality data [4, 3, 24]. Finally, ALD is often underdiagnosed, especially in its early stages when symptoms are mild but intervention is most effective, making improved understanding of ALD onset especially valuable. It is estimated that up to one percent of the population in the United States suffers from ALD [25].

We reconstruct ALD incidence in the United States using mortality data made available by the CDC WONDER database [7]. We relate tallied deaths between 2008 and 2024 to age-structured incidence profiles through well-known disease progression forms and a dynamic backcalculation procedure [26, 27]. We then use a mathematical model to determine the age-structured population and derive age-specific risk as the ratio of incidence to population. Building on these reconstructed forms, we finally develop parameter-free projections for future ALD incidence and risk through 2030. In addition to the overall US population, we provide sex-stratified results.

We find that ALD incidence and the average age at onset have steadily increased since 2008, with a pronounced surge in incidence during the 2020 COVID-19 pandemic [28]. Although upward trends are expected to continue for all age and sex groups, ALD cases are projected to double by 2030 among younger men and older women, highlighting an urgent need for targeted awareness and intervention.

## 2 Model and forecasting overview

The purpose of this work is to estimate ALD risk and incidence in the United States. ALD risk represents an individual’s probability of developing the disease, influenced by personal factors such as alcohol consumption, genetic predisposition, and comorbidities. ALD incidence instead quantifies the population-level occurrence of new cases. Within this framework, ALD incidence density for a given age and year, *I*(*a, t*), is calculated as the product of the estimated age and time specific risk *λ*(*a, t*) of developing ALD, and the corresponding population density *u*(*a, t*), yielding *I*(*a, t*) = *λ*(*a, t*)*u*(*a, t*).

Since the incidence of ALD is difficult to measure directly, we estimate it indirectly using alcohol-related mortality data made available by the CDC WONDER database, where age is grouped into 5-year cohorts [7]. We link deaths to prior disease onset using a known, empirically derived, distribution for the time from ALD onset to death. This distribution allows to construct a mathematical operator that relates observed deaths in a given year to ALD incidence in earlier years. Solving this inverse problem yields estimates of the incidence density *I*(*a, t*), where age is represented as a continuous variable and *t* is the year.

To derive the risk *λ*(*a, t*) we first determine the age-stratified population density *u*(*a, t*) using a mathematical model that incorporates aging, mortality, births, and immigration using data made available by the CDC WONDER database and by the Center for Immigration Studies [7, 29], where age is also grouped into 5-year cohorts. Age-structured population models have been used in conjunction with data assimilation methods to forecast overdose mortality [30, 31], the incidence of chronic disease [27], and are classic tools in demography and epidemiology [32, 33, 34, 35]. In our case, an age-structured model provides data-driven estimates of the population density *u*(*a, t*), where age is treated as a continuous variable following statistical smoothing of the underlying data, and where *t* denotes the year. The risk is then determined as the ratio of the previously estimated incidence and population densities, *λ*(*a, t*) = *I*(*a, t*)*/u*(*a, t*).

Importantly, we enforce smoothness across adjacent age groups within each year for stable estimation, but not across years, allowing incidence and risk to vary over time with possible alcohol-related behavioral changes. The resulting estimate for ALD risk forms a coherent time series across age groups, making it well-suited for data-driven forecasting. To generate short-term projections through 2030 for both ALD risk and incidence, we apply non-negative dynamic mode decomposition (nnDMD), a linear forecasting method that builds on past trends while ensuring all predicted values remain non-negative and biologically plausible. Related mathematical details are provided in Appendix 6. We note that, while the backcalculation procedure uses data through 2024, end-years obtained through backcalculation are generally less reliable in practice; accordingly, we consider reconstructed results only through 2022 [27, 36].

## 3 Data

Population reconstructions were obtained using birth, death, and immigration data made available by the CDC WONDER database [7] and by the Center for Immigration Studies [7, 29]. ALD mortality data were also obtained from the CDC WONDER database, including all deaths listing ALD as an underlying or contributing cause based on the International Classification of Diseases, 10^th^ revision (ICD-10). These causes are: K70.0 (alcoholic fatty liver), K70.1 (alcoholic hepatitis), K70.2 (alcoholic fibrosis and sclerosis of liver), K70.3 (alcoholic cirrhosis of liver), K70.4 (alcoholic hepatic failure), and K70.9 (alcoholic liver disease). The data were stratified by year and sex, and aggregated into 5-year age groups, with suppressed counts due to small numbers treated as zero. We used mortality data from 2008 to 2024 (the last year for which it was made available by the CDC); within this period only a small minority of deaths in the above categories occurred for those younger than 25. The time-to-death distribution for ALD was derived by fitting survival data from a large cohort of ALD patients [37] to a Weibull distribution, commonly used in epidemiological modeling [38, 39, 40]. More details on data sources are provided in Appendix 6.

## 4 Results

We now present our results for age-stratified ALD risk and incidence for 2008-2022, along with projections for 2023-2030; these intervals will hereafter be referred to as the *reconstruction* and *forecast* periods, respectively. Given the structure of our data, we limit reconstruction and forecasting only for individuals aged 25 to 100. Although our backcalculation procedure uses data through 2024, end-year estimates are less reliable in practice, so we focus on reconstructed results through 2022. We first discuss trends for the full population before proceeding to sex-stratified analyses. Since reconstructions and forecasts are performed separately for males, females, and the total population, aggregated values summed across the two sexes do not always match results for the overall population, however, discrepancies are small and findings are in close agreement. Finally, we examine the impacts of a possible intervention scenario, in which awareness campaigns and/or early prevention measures are aimed at one of the most at-risk groups.

### 4.1 Full population

#### 4.1.1 Reconstruction period (2008-2022)

We begin by discussing estimates of ALD incidence for the full population, without age or sex stratification, over the reconstruction period. The total annual incidence is obtained by integrating the age-specific incidence over all ages, 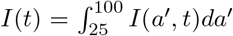. Our results, shown in Fig. 1a with the associated uncertainty, point to significant increases. Incidence rose steadily from about 35,000 to 58,000 cases between 2008 and 2019, a 66% rise. A particularly sharp jump occurred in 2020, coinciding with the onset of the COVID-19 pandemic, with about 109,000 cases, almost double compared to 2019. In 2021, incidence declined to roughly 69,000 cases and further fell to 49,000 in 2022.

**Figure 1:**
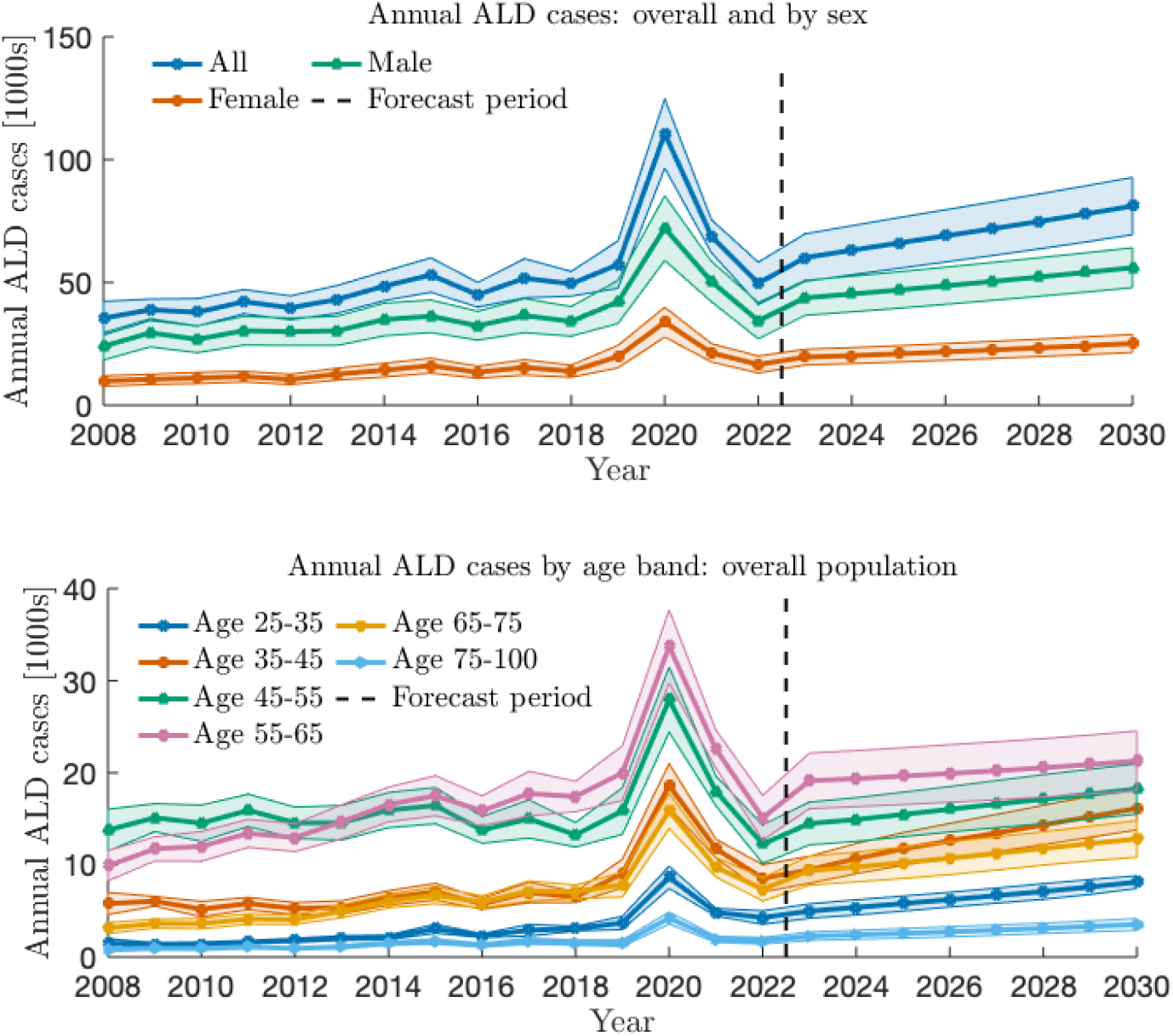
ALD incidence by year in the United States, 2008-2030. The period 2008-2022 contains reconstructed data, the period 2023-2030 contains future forecasts. In Panel (a) data points represent overall ALD incidence per year given by 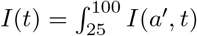 where *I*(*a*′, *t*) is the ALD incidence density obtained via reconstruction. The integral is over all reconstructed ages, from 25 to 100. We include a piecewise linear interpolation between years. The corresponding sex-specific results reveal consistently higher ALD incidence among males. In Panel (b) data points represent ALD annual cases grouped in specific ten-year age-bands given by 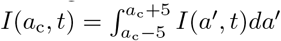 where *a*_c_ = 30, …, 70 are the centers of each age-band, covering ages from 25 to 75. The last age-band groups ages above 75 and defines 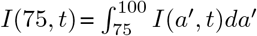. Broadly speaking, ALD incidence rose sharply for all age and sex categories in 2020, concurrent with the onset of the COVID-19 pandemic, and declined in the post-pandemic years. Despite these post-pandemic declines, incidence levels in most groups still surpass pre-pandemic ones, with projections indicating a continued upward trend. Of particular concern is the projected rise in incidence among young adults between 25-35 and 35-45, and among those aged 65-75. Shaded regions indicate uncertainty.

In Fig. 1b we show ALD incidence by age, using ten-year age-bands. Substantial variability across age groups, as well as shifting patterns over time, emerge. The highest ALD incidence is among middle-aged and older adults, particularly those aged 45-55 and 45-65. These groups also exhibited the largest increases in 2020. Interestingly, in 2008, at the beginning of the reconstruction period, ALD incidence was highest among individuals aged 45-55, but by 2013 they were surpassed by those aged 55-65, with the gap between the two age cohorts widening over time. This shift may reflect demographic changes, but also aging of the groups more likely to engage in risky behaviors as discussed below.

Young adults aged 25–35 exhibit the lowest incidence as can be expected since ALD develops cumulatively over time. However, incidence in this group is rising, more than doubling from about 1,500 cases in 2008 to 3,700 in 2019. As for other age groups, incidence spiked sharply in 2020, rising to about 8,700 cases, more than twice 2019 levels, but by 2022 had returned to pre-pandemic levels, at approximately 4,300 annual cases.

Among those aged 35-45, incidence declined or remained stable through the early 2010s, at roughly 6,000 annual cases, and began increasing in 2018, culminating in a three-fold rise to 18,600 cases in 2020. Despite modest subsequent declines, in 2022 incidence remained elevated compared to pre-pandemic levels, at about 8,500 annual cases.

In Fig. 1a we also distinguish between the two sexes. For both sexes near identical temporal trends are observed, consisting of a gradual increase, a sharp spike in 2020, and a modest decline. However, throughout the entire reconstruction period, males exhibited approximately two to three times higher ALD incidence than females.

Given the relationship *I*(*a, t*) = *λ*(*a, t*)*u*(*a, t*) an important question is whether rises in incidence are due to increases in individual risk, measured by *λ*(*a, t*), or in age cohort size, measured by *u*(*a, t*). To better disentangle these effects, and to avoid including possible anomalies introduced by the COVID-19 pandemic, we first focus on the 2008-2019 period. As can be seen from in Fig. 2a, the peak in ALD incidence shifts toward older ages over time. Population trends (not shown) reveal similar age-stratified population increases over the same period, suggesting that cohort size plays a major role in shaping ALD incidence dynamics. However, the estimated risk, shown in Fig. 2b does not remain uniform over time, suggesting that behavioral factors beyond cohort size are also important.

**Figure 2:**
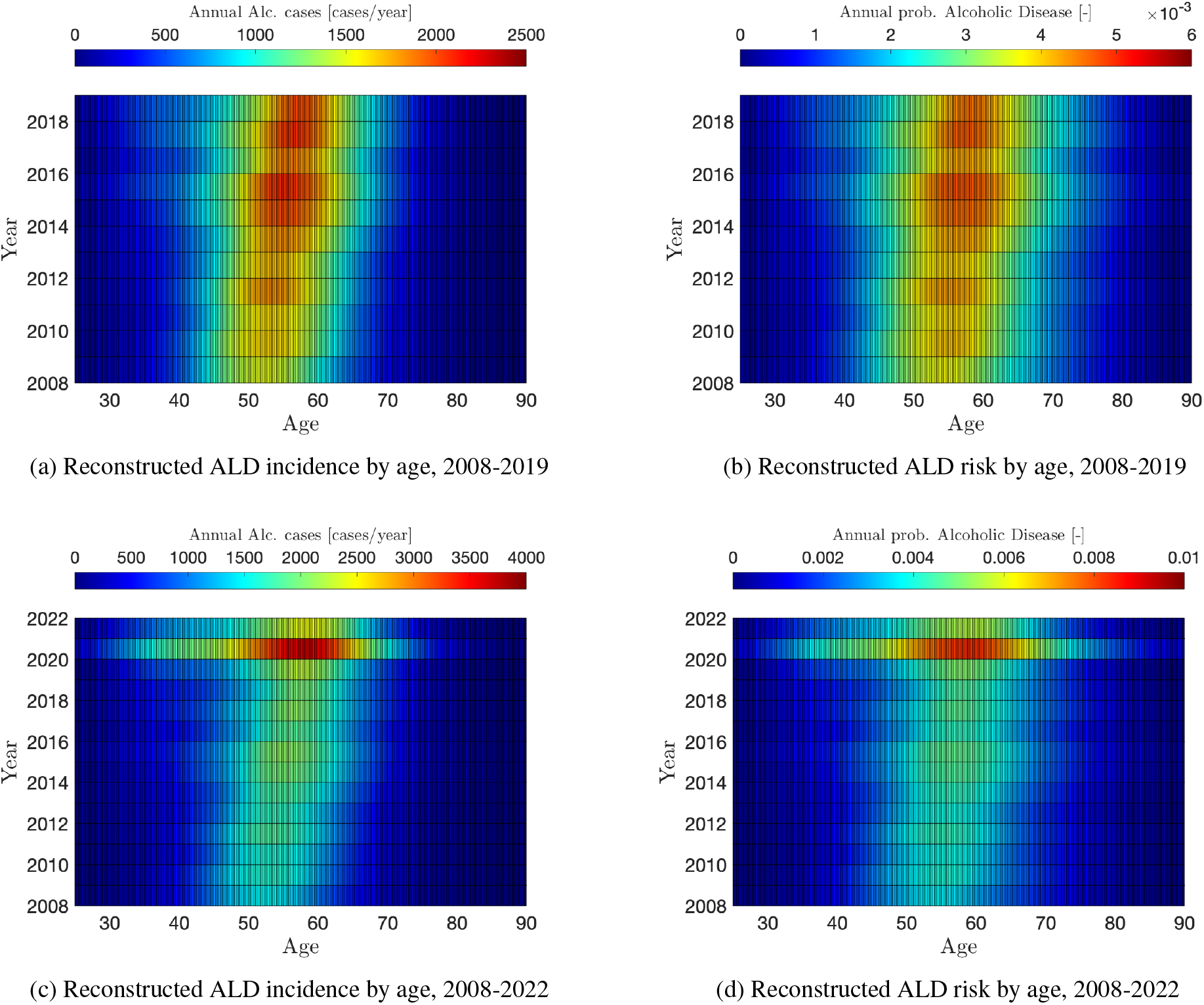
Reconstructed ALD incidence and risk stratified by age. Panels (a) and (b) show ALD incidence and risk over the pre-pandemic period 2008-2019. Over time, the age of highest incidence shifts older, consistent with the aging of large population cohorts. The associated risk, however, is not uniform in time and also shifts older, albeit to a lesser extent. These patterns suggest that cohort effects beyond population size (such as alcohol consumption preferences or other behavioral factors) also contribute to ALD incidence. Panels (c) and (d) show similar trends for ALD incidence and risk over the full reconstruction period (2008-2022). Note however the change in scale and the large increase in incidence and risk for all age groups concurrent with the COVID-19 pandemic in 2020.

Specifically, we observe a general increase and broadening of ALD risk across age groups between 2008 and 2019. In 2008, the 50-55 age cohort had the highest risk of developing ALD, with a likelihood of approximately 2 new annual cases per 1,000. By 2019, the entire 53-66 age range had exceeded this level, with the highest risk concentrated among those aged 55-60. Interestingly, ALD risk decreased among individuals aged 45-55 while increasing among those aged 55-65 over the same period. This finding supports the hypothesis that higher-risk individuals from the 45-55 cohort aged into the 55-65 cohort, and that birth-cohort effects beyond size (such as socioeconomic composition or alcohol-related behavioral factors) also contributed to rising ALD incidence.

Finally, both incidence and risk rose dramatically for all age groups during the COVID-19 pandemic, as shown in Figs. 2c and 2d, where results are plotted for the entire reconstruction period. The scales are much higher than those in Figs. 2a and 2b where only the pre-pandemic period from 2008 to 2019 is considered. Large spikes are observed in 2020 for those aged 45-65; as of 2021, ALD risk remained elevated compared to pre-pandemic levels, particularly for those aged 25-35.

#### 4.1.2 Forecast period (2023-2030)

We now turn our attention to the forecast period. Overall ALD incidence is projected to rise steadily through the end of the decade, rising from the approximately 49,000 cases reconstructed in 2022, to more than 80,000 estimated for 2030, corresponding to a 63% rise. Although these projections remain below the number of cases recorded during the 2020 peak, they are consistent with the upward trend that preceded the pandemic, as shown in Fig. 3a.

**Figure 3:**
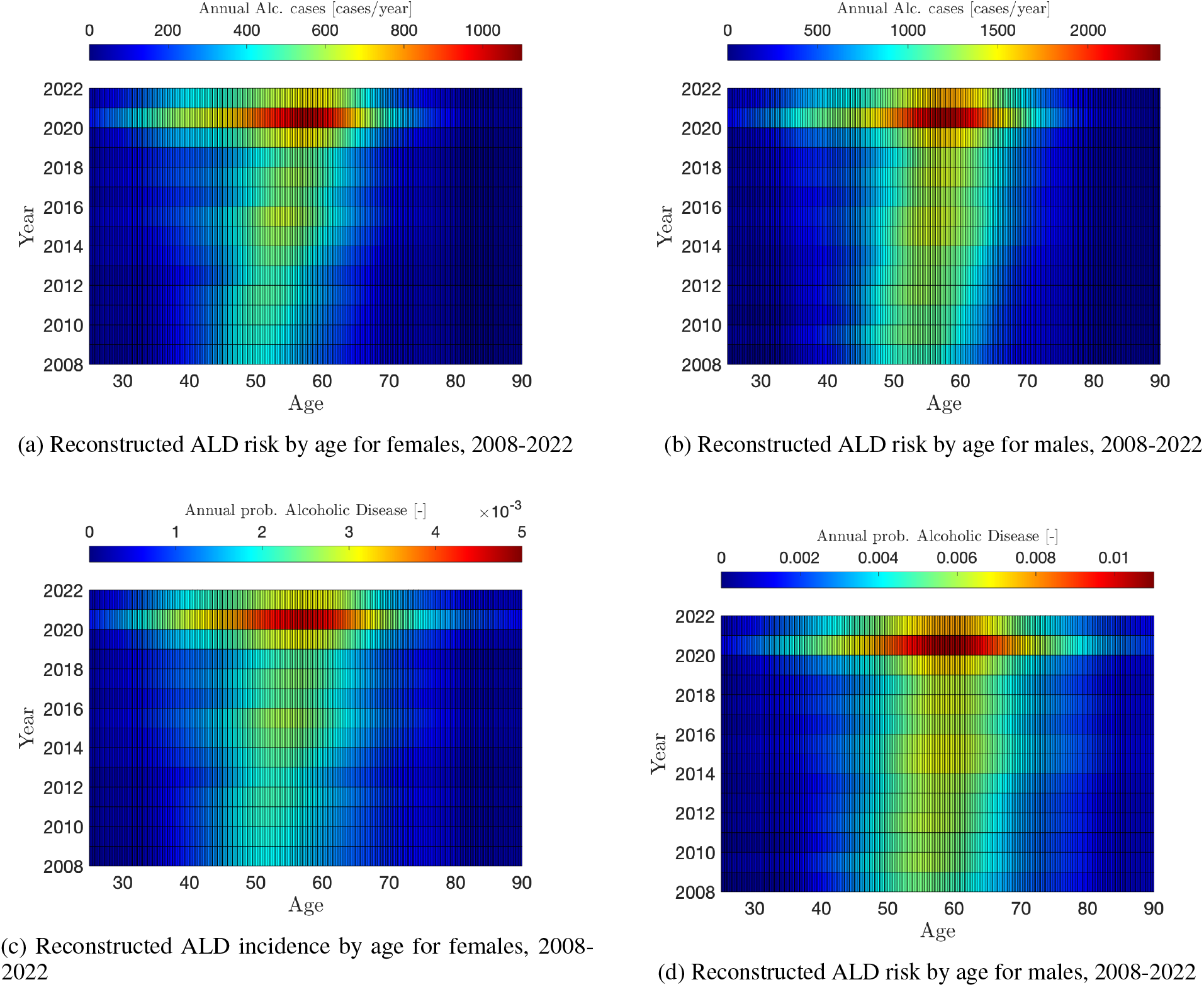
Reconstructed ALD incidence (left) and risk (right) stratified by age. Panels (a) and (b) show ALD incidence and risk over the pre-pandemic period 2008-2019. Over time, the age of highest incidence shifts older, consistent with cohort-size effects as large population cohorts age. The estimated risk, however, is not uniform in time and also shifts towards older ages, albeit to a lesser extent. This pattern suggests that cohort effects beyond population size, including alcohol consumption patterns or other behavioral risk factors may also contribute to ALD incidence. Panels (c) and (d) show ALD incidence and risk over the full 2008-2022 period. Note the change in scale and the large increase in incidence and risk across age groups concurrent with the onset of the COVID-19 pandemic in 2020.

The 55-65 age cohort is expected to remain the highest-incidence group through 2030, with cases increasing by approximately 40% compared to the 2022 reconstructed counts, from about 15,000 to 21,000 cases. The next high-incidence group is expected to be those aged 45-55, for which ALD incidence is expected to increase by about 50%, from 12,000 to 18, 000 in the same time frame. A large increase of about 75% is also expected for those aged 65-55, rising from 7,400 to 13,000 cases.

More concerning patterns emerge among younger adults. For those aged 25-35 and 35-45 incidence is projected to almost double between 2022 and 2030, rising by approximately 90% from 4,300 to 8,000 and 8,500 to 16,000, respectively. Although the number of cases is lowest among those older than 75, we also expect a doubling between 2022 and 2030, from about 1,800 to 3,600 cases.

These increases in ALD incidence will be driven primarily by population growth; risk levels (not shown) are also projected to rise, but to a lesser extent. For example, the largest increase in risk is observed among middle-aged individuals, for whom risk is expected to increase by about 20% between 2022 and 2030.

### 4.2 Sex-specific results

Sex-stratified trends mirror those of the general population, with notable differences between pre- and post-pandemic periods. For both sexes, ALD incidence and risk rose steadily from 2008 to 2019, with a marked surge in 2020; furthermore, for both sexes, the age groups with the highest incidence and risk shifted older over time. Between 2012 and 2019, incidence was higher among males, rising by more than 40% from approximately 30,000 to 43,000 annual cases. Although incidence was lower among females, the number of annual cases almost doubled over the same seven-year period, from approximately 11,000 to 20,000. This relative increase is more than twice the one observed in males, signaling an accelerated rise of ALD among females.

The forecasts shown in Fig. 1a indicate continued increases in overall ALD incidence for both sexes, although at slower overall rates compared to the pre-pandemic years. Between 2022 and 2030, male incidence is projected to rise more than 60%, from approximately 34,000 to 55,000 annual cases, female incidence instead is expected to rise by approximately 50% from 17,000 to 25,000 annual cases.

Our age-stratified analysis reveals that incidence will remain highest among those aged 55-65 and 45-55, and will keep rising for both sexes across all age groups over the entire forecast period, as shown in Fig. 4. In Table 1 we compare the 2022 reconstructed ALD incidence to the 2030 projected number of ALD cases. Within this period, male incidence is projected to double in the 25-35 and 35-45 age groups, with more moderate increases in older age ranges. Among females, incidence is expected to increase more uniformly, though not completely evenly, and at a lower rate than the steepest increases observed in males. From 2022 to 2030 ALD incidence is projected to double among males in the 25-35 and 35-45 age groups, and rise by about 50% and 35 %in the 45-55 and 55-65 agre groups, respectively. Among females, we project growth between 40 and 60% across the four age groups listed above. However, growth rates are expected to be higher for older females than older males: cases will rise by about 70% among females aged 65-75 and will more than double among those older than 75, compared with projected increases of 56% and 85%, respectively, among males in the same age groups. As a consequence of these trends, by 2030, ALD incidence for women aged 45-55 is projected to reach near parity with women aged 55-65, at about 6,000 cases. Notably, female incidence was less dispersed across age groups during the reconstruction period, and is projected to remain so, consistent with prior studies [41, 42, 43].

**Table 1:** Projected growth of ALD incidence in the United States between 2022 and 2030 by age and sex. Reconstructed (2022) and projected (2030) values are rounded to the nearest ten. Significant increases are expected for all categories, especially for males aged 25-35 and 35-45, and females 75 and older, for which cases are expected to double. Prevention and intervention efforts should be prioritized for these groups. Aggregating estimates across age groups yields an overall projected increase of 61% among males and of 51% among females.

| Age group | Males, 2022 | Males, 2030 | Males, pct | Females, 2022 | Females, 2030 | Females, pct |
| --- | --- | --- | --- | --- | --- | --- |
| 25–35 | 2,360 | 5,070 | + 115% | 1,630 | 2,540 | + 56% |
| 35–45 | 5,400 | 10,680 | + 98% | 3,190 | 4,740 | + 48% |
| 45–55 | 8,320 | 12,680 | + 52% | 4,160 | 6,120 | + 47% |
| 55–65 | 10,740 | 14,540 | + 35% | 4,560 | 6,330 | + 39% |
| 65–75 | 5,870 | 9,130 | + 56% | 2,140 | 3,650 | + 71% |
| 75+ | 1,670 | 3,100 | + 85% | 650 | 1,340 | + 106% |

**Figure 4:**
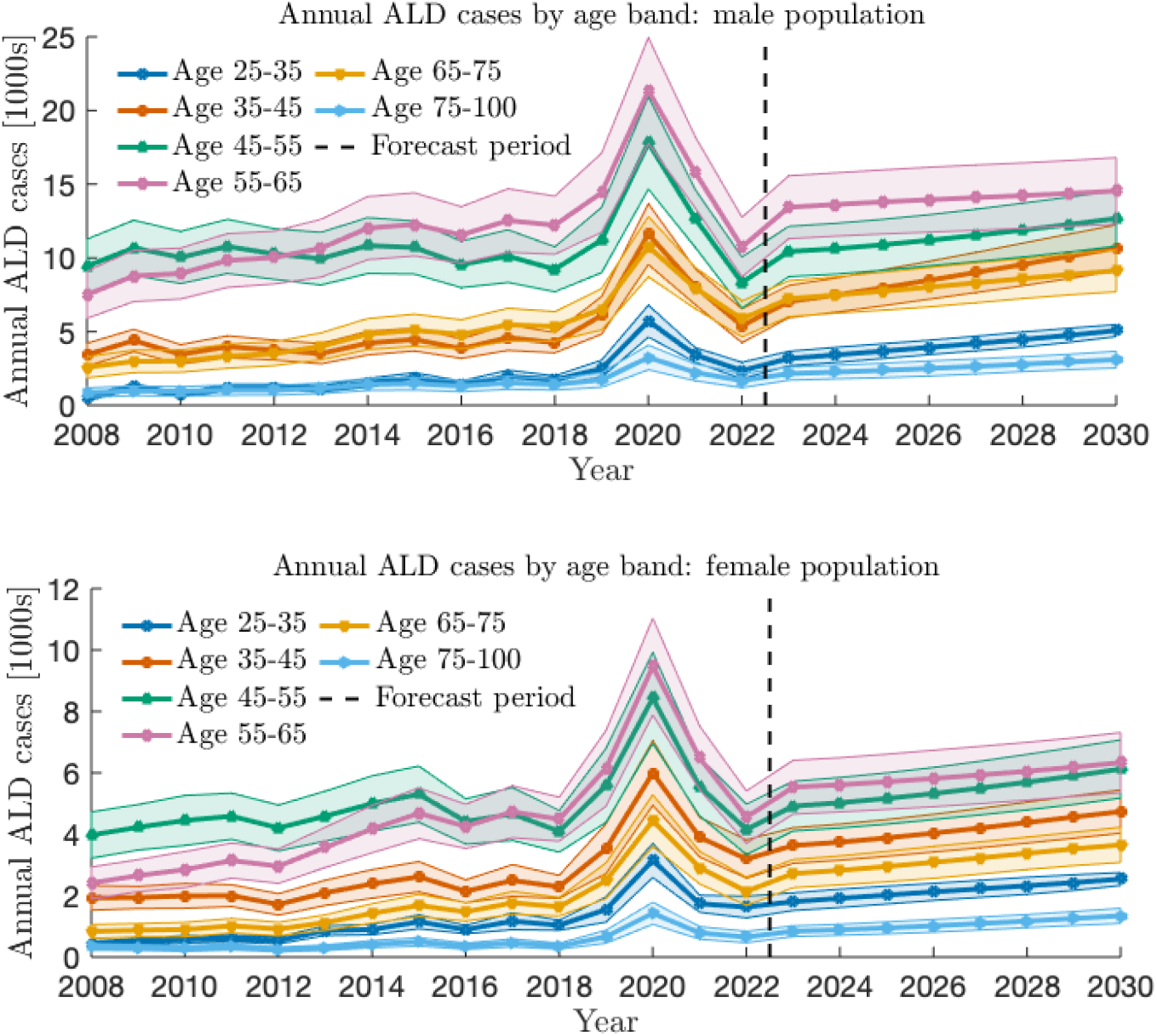
Sex-specific ALD incidence by year and age-band in the United States, 2008-2030. The period 2008-2022 contains reconstructed data, the period 2023-2030 contains future forecasts. The top (lower) panel pertains to the male (female) population. Data points represent ALD annual cases per year grouped in specific ten-year age-bands given by 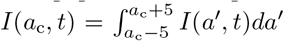 where *I*(*a*′, *t*) is the ALD incidence density obtained via reconstruction, and *a*_c_ = 30, …, 70 are the centers of each age-band, covering ages from 25 to 75. The last age-band groups ages above 75 and defines 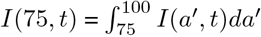. Future ALD incidence is projected to be more than twice as much in males as in females across all age bands. Post-pandemic growth is more heterogeneous across male age groups than female age groups, with particularly large projected increases in younger males aged 25-35 and 35-45, and in older cohorts aged 65-75. Although incidence is projected to grow more uniformly among females, the highest growth is observed in females aged 45–55. Shaded regions indicate uncertainty.

As seen in Fig. 4, without intervention, by 2030 the number of ALD cases among males is projected to be more than double that of females in all age groups. Risk projections (not shown) suggest that the accelerated growth of ALD cases for those aged 25-35 and 35-45 compared with those aged 55-65, in both sexes, is mainly driven by demographic factors, with smaller behavioral risk contributions.

### 4.3 Targeted intervention: younger males

Our forecasts suggest continued growth in ALD incidence in the coming years, particularly among men under 45. This increase is largely independent of population trends, and depends on heightened risk as confirmed by examining the dominant growth modes of the risk-propagation operators described in Appendix 6.3 and plotted in Fig. 6. Given the localized growth dynamics, and the alarming forecasts, we provide a forecast scenario that incorporates interventions prioritizing younger men and show that even modest but targeted approaches can successfully reduce incidence and mitigate future ALD burden. Mathematically, the intervention emerges as an approximate 5% reduction in risk for men aged 30-40. Further details are provided in Appendix 6.4.

Results from the intervention scenario are shown in Fig. 5. We observe reductions in ALD incidence of approximately 55% and 39% in the 25-35 and 35-45 age groups, respectively, in 2030. No substantial changes are observed in other age groups These findings suggest that interventions aimed at reducing alcohol consumption among men under 45 may be a worthwhile, and perhaps necessary, public health measure.

**Figure 5:**
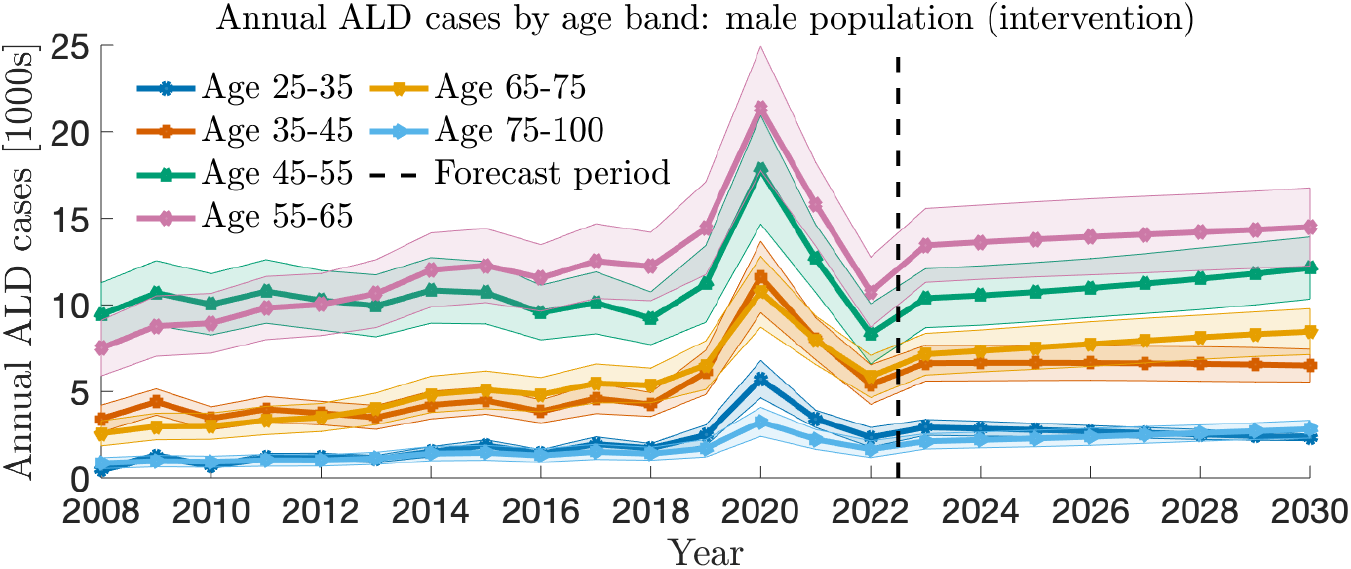
Results of a simulated intervention to reduce risk, targeting younger men between 2022 and 2030. Mathematical details are presented in Appendix 6.3. Roughly speaking, the intervention scenario implemented here can be described as a 5% decrease in risk for younger males. We observe 55% and 39% fewer cases among the 25-35 and 35-45 age groups, respectively, by the end of the simulation period in 2030, showing that even modest intervention can yield noticeable results. Given the target strategy employed, incidence does not change substantially among the groups aged 45 and over.

**Figure 6:**
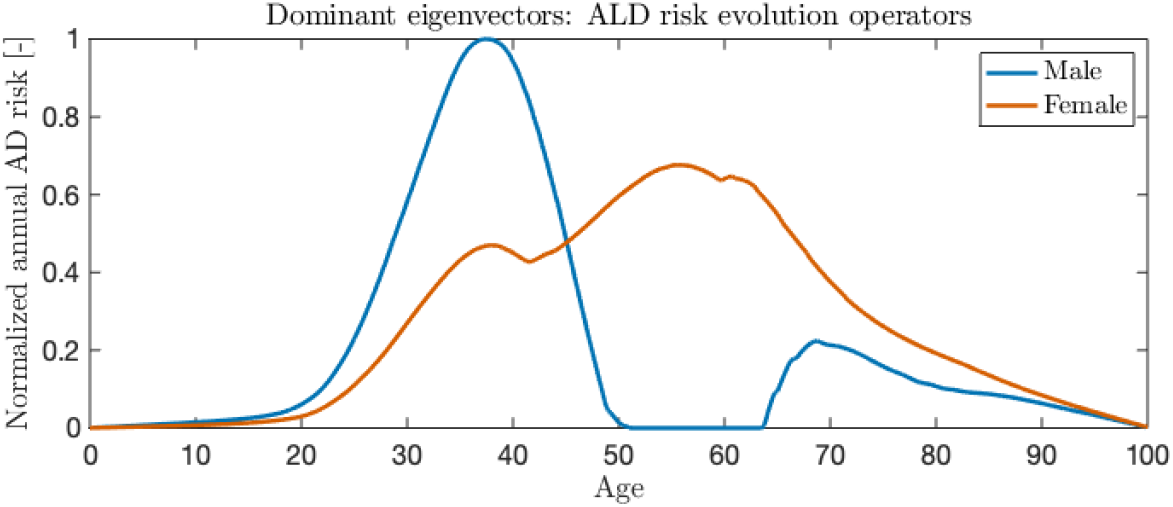
Dominant growth modes for the male and female risk-propagation operators. As our forecasting results suggest, the substantial rise in ALD risk among and middle-aged men between 30 and 40 is largely decoupled from the growth observed in older cohorts. Similar dynamics are not observed among the female population, whose dominant growth mode shows little evidence of the localized, age-specific dynamics observed in males. Our hypothetical intervention scenario focuses on reducing the dominant male eigenvalue corresponding the mode shown here.

## 5 Discussion

Our results reveal a steadily advancing age profile of ALD patients, which began in 2008 (or earlier) and is projected to continue through 2030. Peak ages for both incidence and risk are increasing, reflecting strong cohort effects that go beyond simple population aging. Although incidence will remain highest among those aged 55-65 and 45-55, we project accelerated growth in the 65-75 age group (and for those older than 75) through 2030. This mirrors the structural shift observed in the early 2010s, when the 55-65 age group surpassed the 45-55 age group as the highest-incidence cohort. These projections will require adjustments to the healthcare system, as older ALD patients may have more complex comorbidities, lower physiological resilience, and different support needs than younger individuals.

Our sex-specific analysis further shows that ALD incidence and risk are increasing sharply among males aged 25-35 and 35-45. This suggests a growing need to better understand and address high-risk drinking patterns in younger men to limit future health and societal burdens. Although ALD incidence and risk remain higher among males, growth is sharper among females, who also experience earlier onset of ALD. These patterns emerge directly from our reconstructions, without being structurally imposed, suggesting that they are not artifacts of the modeling framework. This is further supported by the use of separate datasets for overall and sex-specific analyses which yield consistent results.

We also observe spikes in ALD incidence across all sex and age groups in 2020, concurrent with the COVID-19 pandemic. These spikes likely reflect real increases in ALD incidence rather than changes in screening or diagnostic practices, or broader morbidity trends. This interpretation is supported by the use of death counts (based on death certificates) which are less sensitive to diagnostic variation than case counts, and by the fact that ALD was the primary cause of death in most of them. Our reconstruction indicates that by 2022, incidence largely returned to pre-pandemic levels among most age groups of both sexes but began increasing soon thereafter and at different rates depending on the specific demographic group. These findings are consistent with, and extend, prior studies on the accelerated rise of alcohol and drug related mortality and morbidity among older groups, females, and during the pandemic [6, 37, 41, 42, 43, 44, 45, 31, 46, 47].

Our sex-stratified forecasts, valid through 2030, suggest increasing ALD incidence among males aged 25-35 and 35-45, with rates projected to rise and remain above pre-pandemic levels for the foreseeable future. The corresponding rises in females are less pronounced but still significant. These patterns align with the rise in so-called “deaths of despair” among working-age men, which began to be documented in the mid-2010s [48].

While the literature on cohort effects in ALD is limited, our findings agree with one of the few existing relevant studies [20]. Our ALD risk and incidence reconstructions, based on ALD deaths, identify the highest risk among those born in the mid-1950s to mid-1960s, consistent with [20]. More significantly, while [20] further hypothesized strong cohort effects in those born in the mid-to-late 1980s, without stratifying by sex, their data visualization was inconclusive. Our reconstruction confirms this hypothesis, as it reveals elevated ALD risk for men (who account for the majority of cases) aged 35-45, with pronounced cohort effects. Given the strong growth in ALD incidence projected for males aged 35-45 (and 25-35), targeted interventions may be justified. A simulated, modest intervention scenario showed that, roughly speaking, lowering risk by about 5% in these groups could lead to significant reductions in ALD incidence. The relatively young age of this cohort is especially relevant, as cumulative liver damage is more likely to be reversible through behavioral changes and may yield long-term healthcare cost savings [10, 49].

In light of these findings, public health strategies should prioritize targeted prevention and early intervention for high-risk groups. Awareness campaigns should emphasize the cumulative harm from alcohol and the benefits of cessation. Given the rise in ALD incidence during the COVID-19 pandemic, a period of heightened isolation, strong social connections and access to services, may serve as protective factors. Furthermore, the sharper pre-pandemic rise and the slower post-pandemic growth in ALD among females, highlight the need to better understand sex-based differences in adaptation and the contribution of metabolic, mental health and stress factors.

Our study is subject to limitations. As our backcalculation procedure relies on death certificates as its primary input, we implicitly assume that all relevant ALD dynamics are captured by mortality dynamics. While several studies suggest that 5-year ALD survival is poor, and has not improved substantially over time, we acknowledge that these studies are themselves limited and may not fully capture all outcomes. Furthermore, as ALD is often undiagnosed, death certificates almost certainly underestimate true incidence. However, unless underdiagnosis is systematically biased in certain age or sex groups, this primarily affects our reconstructions and forecasts quantitatively, while our qualitative conclusions, including past and future ALD trends, remain valid and useful. Our data-driven forecasting approach extrapolates trends from the training data. Substantial changes in alcohol consumption patterns or the emergence of effective ALD therapies could affect the reliability of our projections. This limitation affects all data-driven and statistical forecasting methods and is not specific to our approach. Furthermore, while our analysis showed the potential for targeted interventions to reduce ALD incidence among younger men, it does not provide any insight on design or implementation. Finally, our computational pipeline offers a consilient approach [50] to addiction research and can be extended by grouping factors such as sex, age, race, and geography to better identify at-risk groups, as well as to other health conditions where age plays an important role.

## Data Availability

CDC WONDER database
Center for Immigration Studies

https://wonder.cdc.gov/

## Acknowledgments

This work was supported by the ARO (grant W911NF-23-1-0129, MRD) and the NSF (grant OAC-2320846, MRD). Moreover, AV and EI were supported by the Indam GNCS (Italian National Group of Scientific Calculus).

## 6 Appendix

In this Appendix we illustrate how to reconstruct both the incidence density *I*(*a, t*) and the risk *λ*(*a, t*), which are not known a priori, using a computational approach that integrates ALD mortality data and mathematical modeling. The procedure is outlined in Section 6.1 whereas parameters and data sources are listed in Section 6.2. Finally, in Section 6.3 we illustrate the methods used to obtain future projections for ALD incidence and risk. All quantities are considered in the context of the population of the United States.

### 6.1 Demographic model

We begin by defining the ALD incidence density *I*(*a, t*) as the density of new ALD cases among individuals of age *a* at time *t*, given by

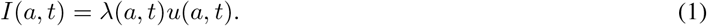

Accordingly, the number of new ALD cases among individuals in the age interval [*a, a* + *da*) at time *t* is *I*(*a, t*)*da*. Here, *u*(*a, t*) denotes the population density of persons of age *a* at time *t* and *λ*(*a, t*) is the ALD risk hazard, defined as the probability of developing ALD at age *a* and time *t*, conditional on not having developed ALD previously. Neither *λ*(*a, t*) nor *u*(*a, t*) are known a priori.

To estimate *u*(*a, t*) we use an age-structured McKendrick model [51, 52, 53], a classical framework in population dynamics describing the evolution of age distributions over time. We write

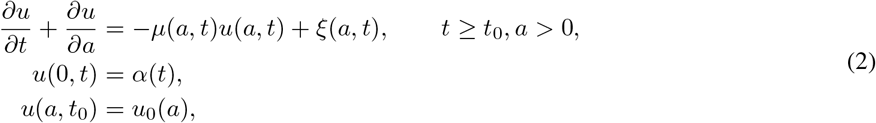

where *µ*(*a, t*) *>* 0 is the mortality hazard rate at age *a* and at time *t*, and *ξ*(*a, t*) ≥ 0 is the rate density such that *ξ*(*a, t*)*da* yields the rate at which individuals with ages [*a, a* + *da*) immigrate into the population at time *t*. The age boundary condition *u*(0, *t*) represents the inflow of newborns, given by the birth rate *α*(*t*) at time *t*. Finally, *t*_0_ denotes the initial time, set to 2008, and *u*_0_(*a*) is the corresponding initial population density distribution at time *t*_0_, which is assumed to be known. In this setting, the number of individuals with ages between [*a, a* + *da*) at time *t* is given by *u*(*a, t*)*da*, the probability that an individual aged *a* at time *t* dies during the time interval [*t, t* + *dt*) is *µ*(*a, t*)*dt*, and the number of deaths for individuals of ages between [*a, a* + *da*) occurring in the interval [*t, t* + *dt*) is their product *µ*(*a, t*)*u*(*a, t*)*da dt*. The quantities *µ*(*a, t*), *ξ*(*a, t*), *α*(*t*), *u*_0_(*a*) are assumed to be known and discussed in Section 6.2. To make progress, once *u*(*a, t*) is derived from Eq. 2, we discretize the age domain *A* = [*a*_min_, *a*_max_) into *K* non-overlapping intervals *A*_*k*_ so that

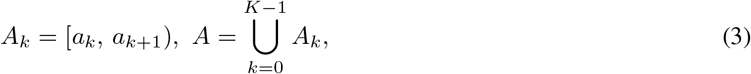

where *a*_0_ = 0 and *a*_*K−*1_ is the left endpoint of the final interval, i.e. *A*_*k−*1_ = [*a*_*K−*1_, *a*_*max*_ = 100). Within this framework, the number of alcohol-related deaths *d*_*kj*_ in an age interval *A*_*k*_ at a time *t*_*j*_ is given by

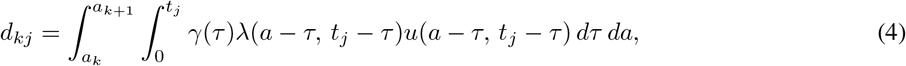

where *γ*(*τ* )*dτ* is the rate of alcohol-related death at time *τ* after onset of ALD and *λ*(*a* − *τ, t*_*j*_ − *τ* )*u*(*a* − *τ, t*_*j*_ − *τ* ) is the incidence density at time *t*_*j*_ − *τ* for individuals of age *a*. Equation 4 implies that the number of deaths observed at time *t*_*j*_ for individuals in the *A*_*k*_ cohort is associated with ALD incidence occurring at time *t*_*j*_ − *τ* and that the time-to-death after disease onset is *τ* . In general *τ* may vary, but the total elapsed time from incidence to death must sum to *t*_*j*_. Furthermore, Eq. 4 allows us to link the continuous age-time formulation for *u*(*a, t*) with discretely observed data aggregated over age intervals *A*_*k*_ and years 2008 ≤ *j* ≤ 2024. The age limits *a*_min_, *a*_max_, the number of deaths *d*_*kj*_, and the form of *γ*(*τ* ) are assumed to be known and discussed in Section 6.2.

To finally reconstruct the risk *λ*(*a, t*) we define the following piecewise constant basis for the age-time domain:

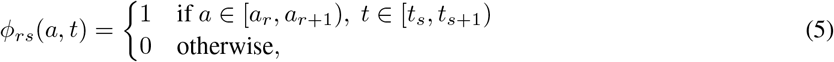

and further assume

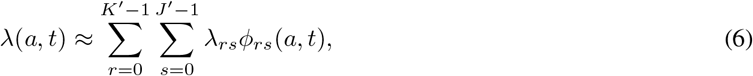

where *λ*_*rs*_ are unknown. Substituting Eq. 6 into Eq. 4 yields:

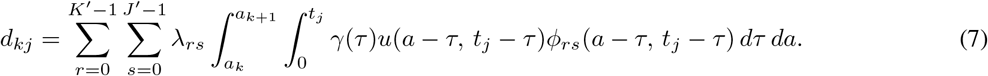

The *K*′, *J*′ partitions imposed on the discretization of *λ*(*a, t*) are chosen to ensure numerical stability and sufficient resolution. Following the procedures shown in [27, 26], and starting from Eq. 7, we obtain the discretized system

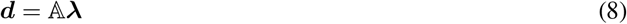

where ***d*** ∈ ℝ^*K*·*J*^ is the vector of observed ALD deaths in age cohort *k* at year *j* of components *d*_*kj*_, ***λ*** ∈ ℝ^*K′*·*J′*^ is the vector of risk components *λ*_*rs*_ to be estimated, and 𝔸 ∈ ℝ^*K*·*J×K′*·*J*′^ is the discretized operator:

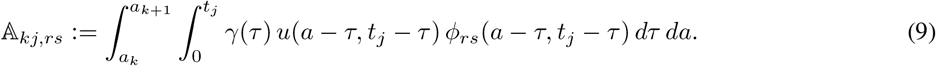

Note that 𝔸 is underdetermined in general, hence solving the linear system in Eq. 8 requires additional regularization. In practice, we add a Laplacian-based Tikhinov regularization within each year to ensure within-year smoothness in the reconstruction [54]; however, we do not enforce any smoothness conditions across years, as we expect that year-to-year risk may change across age cohorts due to demographic, environmental, or other factors. In [27], this formulation was proven to be well-posed. Independent reconstructions were performed for the overall, male, and female populations.

### 6.2 Data sources and model parameters

Here, we describe the sources of our data and the modeling choices made in our backcalculation reconstructions of *λ*(*a, t*) and *I*(*a, t*). The required quantities are *µ*(*a, t*), *ξ*(*a, t*), *α*(*t*), *u*_0_(*a*), *d*_*kj*_, *γ*(*τ* ), *K, J*; how we obtained them is discussed below. The initial year is set at *t*_0_ = 2008 and we carried out reconstructions until 2024. As discussed in the main text however, we analyze the reconstructed data only through 2022 since backcalculation results for the first and final years are generally less reliable than for those in the middle years. Early-year estimates lack information from preceding data and are more sensitive to initial conditions, while final-year estimates are less reliable due to reporting delays and limited future information to inform the backcalculation itself [27, 36].

#### 6.2.1 Population construction

We obtained birth, death, and demographic data from the CDC WONDER database and constructed *µ*(*a, t*), *u*_0_(*a*) following the approach described in [26, 27]. Briefly, we obtained *µ*(*a, t*) through an ensemble Kalman filter inversion, assimilating observed death data into a dynamic PDE model for the population density *u*(*a, t*). The initial population distribution *u*_0_(*a*) was generated as an ensemble of 1,000 population densities, defined such that their distribution across each 5-year age group was consistent with 2000 census data; within each discrete age group, the population structure was randomly generated following a uniform distribution. Complete details can be found in [26]. The immigration rate density *ξ*(*a, t*) was obtained by fitting a Weibull distribution to yearly data obtained from the Center for Immigration Studies [7, 29]. The Weibull distribution was chosen as its shape and scale parameters showed good empirical fit with the available data. We discuss its features more in detail in 6.2.4. Finally, we note that the last available year for immigration data was 2019; for all future years (in both reconstruction and forecast periods), we thus assumed a continuation of 2019 immigration levels. Annual births *α*(*t*) are directly available from the CDC WONDER database, and injected uniformly throughout the year into the model for the reconstruction period.

#### 6.2.2 Uncertainty quantification

As mentioned in the preceding section, the simulations were performed over an ensemble of 1,000 population distributions. The point estimates reported in the main text correspond to the ensemble mean of the resulting distribution, with the reported uncertainty corresponding to the 5% and 95% limits of the ensemble distribution.

#### 6.2.3 Mortality data

Mortality data were compiled by the National Vital Statistics System and made publicly available by the CDC WONDER database [7]. Final death tallies are available only through the year 2024. We considered all death certificates in which ALD was listed as a cause of death (contributing or underlying) according to the International Classification of Diseases 10th revision (ICD-10). These are: K70.0 (alcoholic fatty liver), K70.1 (alcoholic hepatitis), K70.2 (alcoholic fibrosis and sclerosis of liver), K70.3 (alcoholic cirrhosis of liver), K70.4 (alcoholic hepatic failure), and K70.9 (alcoholic liver disease). The data were further stratified by year, sex, and 5-year age group. We note, however, that we consider <1 year, 1-4 years, and 100+ as distinct age brackets, with all other ages following the standard 5-year bracket structure, giving *K* = 22 total age cohorts. In some instances, data were suppressed due to small numbers; these were considered to be zero. With these choices, the *d*_*kj*_ values that appear in Eq. 4 are all deaths reported by the CDC WONDER database in the categories listed above for years *j* ∈ {2008, …, 2024} and in the *k*^th^ age cohort. Finally, we set *J* = 400, corresponding to a resolution of 0.25 years in the age domain.

#### 6.2.4 Time-to-death distribution *γ*(*τ* )

We obtained the time-to-death distribution *γ*(*τ* ) by fitting empirical survival data among diagnosed ALD patients as reported in [37]. We used a Weibull distribution, which is commonly used to model lifetimes and failure rates, given by

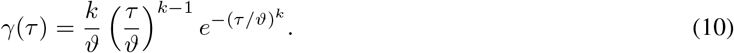

Here, the shape parameter *k* controls failure patterns: *k <* 1 implies failures decrease over time (early failures are more common) and *k >* 1 implies failures increase over time (failures are more likely as time passes due to deterioration or wear-out). The scale parameter *ϑ* controls the time scale of the distribution, with larger *ϑ* values indicating that failures occur at later times. The data we fit Eq. 10 to came from a large-scale cohort study on 11,873 persons which found a 37% five-year survival rate among ALD patients, with 65% of deaths directly due to ALD, and ALD/alcohol use a contributing factor in the remaining 35% of cases [37]. Using a Gauss-Newton nonlinear least-squares algorithm, we found *θ* ≈ 4.96 and *k* ≈ 0.499. Using these parameter estimates, the fitted distribution in Eq. 10 exhibits a 1.9% discrepancy relative to the empirical data, as quantified by the mean absolute percentage error

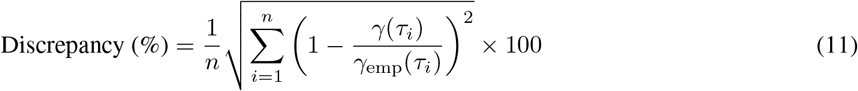

where *n* is the number of observations, *τ*_*i*_ denotes the time of the *i*-th data point, *γ*_emp_(*τ*_*i*_) is the reported survival at time *τ*_*i*_, and *γ*(*τ*_*i*_) is the corresponding value predicted by the fitted distribution in Eq. 10.

#### 6.2.5 Visualizations

Our reconstructions yield continuous forms for ALD density incidence *I*(*a, t*). To visualize the overall number of cases *I*(*t*) in year *t*, as shown in Fig. 3, we plot the integrated quantity

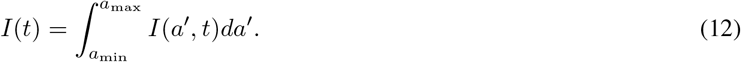

ALD incidence in the ten-year age-bands, as shown in Figs. 3 and 4, are instead obtained as

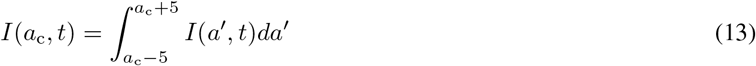

where *a*_c_ = 30, …, 70 are the centers of each ten-year age-band between ages 25 and 75. The last age-band group in Figs. 3 and 4 aggregates all ages above 75 and corresponds to

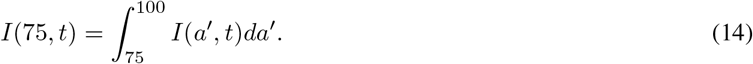

Note that the CDC WONDER database reports data in five-year age cohorts, which we used to reconstruct the continuous incidence density *I*(*a, t*). For the purposes of visualization, we aggregate the derived *I*(*a, t*) into ten-year age-bands centered on the *a*_c_ values listed above, although alternative age groupings could also have been used. Finally, incidence counts are rounded to the nearest 1,000 for larger values and to the nearest 100 for smaller ones (typically, for age-stratified data), unless otherwise specified.

### 6.3 Projection into future years

In addition to providing estimates of past incidence and risk, our reconstructed risk hazard *λ*(*a, t*) forms a time series with substantial coherence across age-bands over time. As such, it is well-suited for purely data-driven, unsupervised machine learning methods for short-term future extrapolation. To do this, we use non-negative dynamic mode decomposition (nnDMD), first introduced in [26] and generate forecasts for both ALD risk *λ*(*a, t*) through 2030. We thus set

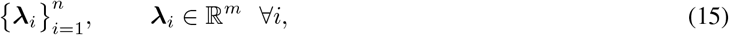

where *n* is the number of observations, and *m* the dimension of each observation. We arrange these snapshots into two *m × n* − 1 matrices Λ_1_, Λ_2_ column-wise as follows:

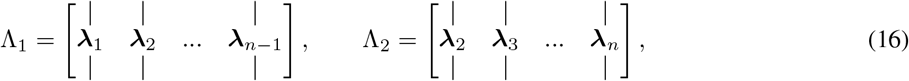

assuming, in this case, *n* ≪ *m*. We seek to reconstruct the operator *K* mapping Λ_1_ to Λ_2_, that is:

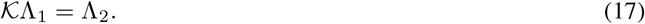

The standard DMD algorithm [55, 56] finds:

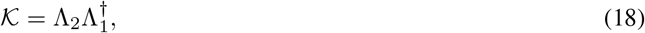

where 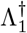 denotes the pseudoinverse. However, as (18) is a purely data-driven regression on an underdetermined problem, it often does not perform well outside of its training interval (or in some cases, even within the training interval). Several modifications of DMD have shown, in different ways, that incorporating problem-relevant information can greatly improve DMD results, particularly for forecasting purposes [57, 58, 59, 60]. The nnDMD formulation used herein instead solves the problem:

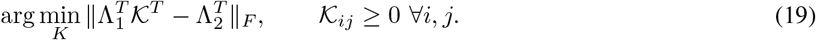

where ∥ · ∥_*F*_ denotes the Frobenius norm. The approach (19) was shown in [26] to provide physically consistent, plausible forecasting trends when applied to similar data as in the present; we refer interested readers to that work for further details on nnDMD, including numerical and theoretical aspects.

Once projections for *λ*(*a, t*) are derived we can adapt the procedure described above to also project the general population mortality *µ*(*a, t*) and extrapolate further estimates of *u*(*a, t*) via Eq. 2. This procedure allows us to also forecast ALD incidence density *I*(*a, t*) = *λ*(*a, t*)*u*(*a, t*) throughout 2030. We did not project immigration rates and used the last available values of *ξ*(*a, t*).

### 6.4 Mathematical details: targeted intervention for young males

Here we provide further mathematical details on the intervention scenario described in the main text. To quantify the potential effect of a targeted intervention, we construct a counterfactual scenario in which the excess growth specific to young males is neutralized. Let 𝒦 ∈ ℝ^*m×m*^ denote the non-negative risk-propagation operator obtained via nnDMD for the male population, with eigendecomposition

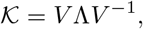

where Λ = diag(*λ*_1_, *λ*_2_, …, *λ*_*m*_) contains the eigenvalues ordered by decreasing magnitude and *V* is the matrix of corresponding right eigenvectors. The dominant eigenvalue satisfies *λ*_1_ ≈ 1.044, with its associated eigenvector strongly localized on the age interval [25, 45] as shown in Fig. 6.

For the intervention scenario, we define the modified operator:

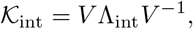

where:

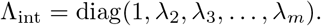

That is, only the dominant eigenvalue is reset to 1 (neutral growth), while all other eigenvalues and the complete set of eigenvectors remain unchanged. This corresponds to a 4.2% decrease in *λ*_1_, which in the main text is referred to as a roughly 5% reduction in risk for younger men. The modified operator 𝒦_int_ is then used in place of 𝒦 to propagate the risk vector ***λ*** from 2022 (the last reconstruction year we assumed to be reliable) forward, exactly as described in Appendix 6.3.

Because the intervention acts solely on the heavily-localized dominant male growth mode, as shown in Fig. 5, risk and incidence trajectories in all other age groups are largely unaffected. For the targeted young male cohorts, the procedure yields substantial reductions. As discussed in the main text, by 2030, projected ALD incidence in the 25–35 age group is approximately 55% lower than in the baseline forecast, while incidence in the 35–45 age group decreases by approximately 39% as shown in Fig. 5 of the main text.

